# “GrimAge,” an epigenetic predictor of mortality, is accelerated in Major Depressive Disorder

**DOI:** 10.1101/2020.12.25.20248290

**Authors:** Ekaterina Protsenko, Ruoting Yang, Brent Nier, Victor Reus, Rasha Hammamieh, Ryan Rampersaud, Gwyneth W. Y. Wu, Christina M. Hough, Elissa Epel, Aric Prather, Marti Jett, Aarti Gautam, Synthia H. Mellon, Owen M. Wolkowitz

**Author notes:** Ekaterina Protsenko, University of California San Francisco School of Medicine.

## Abstract

Major Depressive Disorder (MDD) is associated with premature mortality and is an independent risk factor for a broad range of diseases, especially those associated with aging, such as cardiovascular disease, diabetes, and Alzheimer’s Disease. However, the pathophysiology underlying increased rates of somatic disease in MDD remains unknown. It has been proposed that MDD represents a state of accelerated cellular aging, and several measures of cellular aging have been developed in recent years. Among such metrics, estimators of biological age based on predictable age-related patterns of DNA methylation (DNAm), so called ‘epigenetic clocks’, have shown particular promise for their ability to capture accelerated aging in psychiatric disease. The recently developed DNAm metric known as ‘GrimAge’ is unique in that it was trained on time-to-death data and has outperformed its predecessors in predicting both morbidity and mortality. Yet, GrimAge has not been investigated in MDD. Here we measured GrimAge in 49 somatically healthy unmedicated individuals with MDD and 60 age-matched healthy controls. We found that individuals with MDD exhibited significantly greater GrimAge relative to their chronological age (‘AgeAccelGrim’) compared to healthy controls (p=0.001), with a median of two years of excess cellular aging. This difference remained significant after controlling for sex, current smoking status and body-mass index (p=0.015). These findings are consistent with prior suggestions of accelerated cellular aging in MDD, but are the first to demonstrate this with an epigenetic metric predictive of premature mortality.

## Introduction

Major depressive disorder (MDD), the leading cause of disability worldwide(1), is associated with early mortality(2) and is an independent risk factor for a variety of diseases associated with the aging process(3), such as cardiovascular disease(4–7), dementia(8), osteoporosis(9), and diabetes(10), among others. Even after accounting for lifestyle factors, MDD remains an independent risk factor, raising the possibility of an underlying mechanism of accelerated biological or cellular aging(3,11).

Recently, epigenetic age has emerged as an especially promising measure of cellular aging that may show stronger associations with mortality than earlier metrics of biological age, such as leukocyte telomere length(12). It is based on the finding that chronological age has predictable effects on DNA methylation patterns at subsets of the 28 million 5’-C-phosphate-G-3’ (CpG) sites scattered throughout the genome(13). ‘Epigenetic clocks’ consist of small sets of these CpGs whose methylation patterns yield an estimate of biological or cellular age (‘DNAm Age’), as opposed to strictly chronological age. The specific selections of CpGs vary between the available ‘clocks,’ and are a function of what data the machine learning-derived metric was trained on. When an individual’s ‘DNAm Age’ exceeds their chronological age, they are said to experience ‘Epigenetic Age Acceleration.’ However, most measures of ‘DNAm Age’ and ‘DNAm Age Acceleration’ are only modest predictors of mortality, likely because their derivation was based on chronological age, which by its definition excluded those CpG sites that would signal a departure from normal aging(14).

In an effort to improve upon mortality prediction, the ‘GrimAge’ clock used a two-stage approach to derive its algorithm(15). In the first stage, CpG sites were identified whose methylation states closely correlated with either serum protein levels that predicted mortality, or with self-reported smoking history. In the second stage, these ‘DNAm surrogates’ for serum proteins and smoking history were further trained on large-scale time-to-death data. Not surprisingly, GrimAge outperformed its predecessors in its ability to predict mortality in five independent cohorts(15). It was also able to predict time to onset of coronary heart disease, and has shown associations with congestive health failure, hypertension, type-2 diabetes, physical functioning, comorbidity, and early menopause(15). In all, because GrimAge’s component CpG sites are surrogates for health- and disease-related proteins, as well as smoking history, it demonstrates superior associations with all-cause mortality and age-related health status(16), and is therefore likely to be more sensitive for accelerated cellular aging in psychiatric conditions than its predecessors.

Despite the consensus that depression is associated with premature morbidity and mortality, there have been very few published investigations into potential changes in epigenetic aging in MDD(17), and none using the GrimAge metric. In light of the relative dearth of evidence on epigenetic aging in MDD, we investigated GrimAge Acceleration (‘AgeAccelGrim’) in a cohort of somatically healthy, unmedicated individuals with moderate-to-severe MDD, compared to a group of medically and psychiatrically healthy controls. We hypothesized, based on evidence of early mortality among depressed individuals, that GrimAge Acceleration would be greater in MDD than in similarly aged healthy controls.

## Methods

### Ethics statement

The University of California, San Francisco (UCSF) Institutional Review Board (IRB) approved the study protocol. All study participants gave written informed consent to participate in this study and were compensated for participating.

### Recruitment procedures and study participants

MDD (n⍰ =⍰50) outpatients and healthy controls (n⍰= ⍰63) were recruited by flyers, Craigslist postings, newspaper ads and, in the case of MDD subjects, clinical referrals. All diagnoses, including MDD, were made according to DSM-IV guidelines(18), which were in use at the start of this study. Diagnoses were established using the Structured Clinical Interview for DSM-IV TR(19) and verified in a separate unstructured diagnostic evaluation by a Board-certified psychiatrist. Depression symptom severity was assessed in MDD subjects using the Hamilton Depression Rating Scale (HDRS)(20), while depression severity across MDD and control participants was assessed with the self-rated Inventory of Depressive Symptoms (IDS-SR)(21). All MDD subjects had a minimum 17-item HDRS score of 17. MDD subjects were excluded if they met DSM-IV criteria for any of the following: (i) bipolar disorder, (ii) alcohol or substance abuse within the preceding 6 months, (iii) PTSD or an eating disorder within 1 month of entering the study, and (iv) for any history of psychosis outside of a major depressive episode, or the presence of any psychotic symptoms during the current major depressive episode. Potential healthy controls were excluded for any history of DSM-IV Axis-I diagnoses. All study participants were free of chronic illnesses or acute illnesses or infections, inflammatory disorders, neurological disorders, or any other medical conditions considered to be potentially confounding, as assessed by history, physical examinations, and routine blood screening. All subjects were free of psychotropic medications (including antidepressants), hormone supplements, steroid-containing birth control or other potentially interfering medications, including vitamin supplements above the U.S. recommended daily allowances (e.g. >90⍰mg/day for Vitamin C) and had not had any vaccinations for at least 6 weeks prior to enrollment. For MDD subjects, short-acting sedative-hypnotics were allowed as needed up to a maximum of three times per week, but none within 1 week prior to blood draws in the study. All subjects had to pass a urine toxicology screen for drugs of abuse and a urine test for pregnancy in women of child-bearing age on the day of blood draw.

### DNA Preparation and analysis of methylation

Blood samples were drawn in the morning following an overnight fast. Whole blood was collected in acid citrate dextrose (ACD) tubes for preparation of DNA. Aliquotted samples were stored frozen at -80° until use. DNA was extracted from whole blood using QIAmp DNA purification kits (Qiagen, Redwood City, CA), followed by quality check using a Tapestation (Agilent). Identification and analysis of methylated CpGs used protocols used by our group previously(22). Genomic DNA (500⍰ng) was treated with sodium bisulfite using the Zymo EZ96 DNA Methylation Kit (Zymo Research, Orange, CA, USA), and genome-wide DNA methylation patterns were profiled using the Infinium HumanMethylation450 BeadChip array (Illumina, Inc., San Diego, CA, USA). BeadChips were washed, single-base extension labeled and stained with multiple layers of fluorescence followed by scanning using the Illumina iScan system (Illumina Inc, CA). The samples were randomized on plates. No visual batch effects were found between the plates. IDAT files containing the raw intensity signals were generated using Illumina’s iControl software. Noob background correction(23) was used to pre-process the data prior to submitting it to the DNAm age website https://dnamage.genetics.ucla.edu for analysis. A principal component analysis (PCA) was conducted on Noob-corrected data to identify outliers with DNA methylation assay anomalies. Three healthy controls and one MDD were excluded for the quality control. All the samples were processed and assayed together, and PCA analysis confirmed no significant batch effect existed on experimental blood collection batches. All samples passed quality control, and mean absolute difference between samples and the “gold standard” (which is the mean of a large whole-blood DNAm cohort) were all lower than 0.08.

### Epigenetic Clocks

The GrimAge clock has been previously described(15). ‘GrimAge Acceleration’ is defined as the residual from regressing epigenetic age on chronological age, and is denoted by the prefix “AgeAccel” (e.g., “AgeAccelGrim”). The GrimAge clock is constructed as the composite of 8 DNA methylation-based markers for plasma proteins and self-reported smoking packyears(15). The plasma protein surrogates include: cystatin C, leptin, tissue inhibitor metalloproteinases 1 (TIMP1), adrenomedullin (ADM), beta-2-microglobulin (B2M), growth differentiation factor 15 (GDF15), and plasminogen activation inhibitor 1 (PAI-1). The rationale for selection of these proteins, and their functions and disease-associations are described in Lu et al., 2019 (**see ref. (15)**). The DNAm surrogates for these proteins and smoking history are denoted by the prefix “DNAm” (e.g., “DNAmPACKYRS” for the surrogate of smoking history), and the residuals from regressing on chronological age are specified as “age-adjusted.”

### Statistical Methods

The sample size included was based on all subjects with available data from the parent study (“Cell Aging in Major Depression;” R01 MH083784). The resulting sample was sufficient to detect effect sizes for independent sample t-tests of 0.54 or greater, with a power of 0.80 and a two-tailed alpha= 0.05.

P values for all analyses reflect two-tailed significance with an alpha=0.05. Correlations between epigenetic age and chronological age were based on Spearman correlations. All measures of epigenetic age were subsequently regressed on chronological age, and the residuals were used for all ANCOVA analyses. The age-adjusted residual of GrimAge is referred to as “AgeAccelGrim” to maintain consistency with prior literature. All variables were checked for normality by a Shapiro-Wilk test, and Blom-transformation(24) was used to achieve normality for AgeAccelGrim. Several of the age-adjusted DNAm surrogate marker components of GrimAge were also Blom-transformed: DNAmCystatinC, DNAmLeptin, DNAmPACKYRS, and DNAmTIMP1. For all sub-cohort analyses, Blom transformation was repeated within the sub-cohort prior to analysis. The remaining age-adjusted DNAm components were normally distributed.

All group differences in age-adjusted epigenetic age metrics were initially tested using independent samples T-tests (‘**Model 1**’). Since one of the DNAm surrogate markers included in the GrimAge algorithm was derived nominally as a surrogate for lifetime pack-years of smoking (DNAmPACKYRS)(15), we covaried for current tobacco use in a separate ANCOVA (‘**Model 2**’). We additionally covaried for sex and BMI in ‘**Model 3**,’ as both have been repeatedly shown to modify measures of epigenetic aging(15,25–28). The same models were applied to the individual GrimAge surrogate marker components.

To control for any lingering effects of smoking not accounted for by covariance, we completed sensitivity analyses within a sub-cohort of ‘current non-smokers’ (37 MDD, 55 HC; ‘**Model 4-5’ in Table 3**). While ‘current’ smoking status was used in the primary analyses to maximize sample size, additional data on lifetime smoking exposure were available in a subset of participants (41 MDD, 51 HC). Models 2 and 3 for AgeAccelGrim and DNAmPACKYRS were repeated in this subset to ensure the effect did not differ when lifetime smoking was accounted for, defined as ‘never,’ ‘former,’ or ‘current’ smoker (‘**Model 6-7’ in Supplementary Table 1**). Finally, we excluded the DNAmPACKYRS component from AgeAccelGrim, as has been previously described (22). These analyses are further detailed in the **Supplementary Materials**, except Models 4-5 which are detailed below.

**Table 1:**
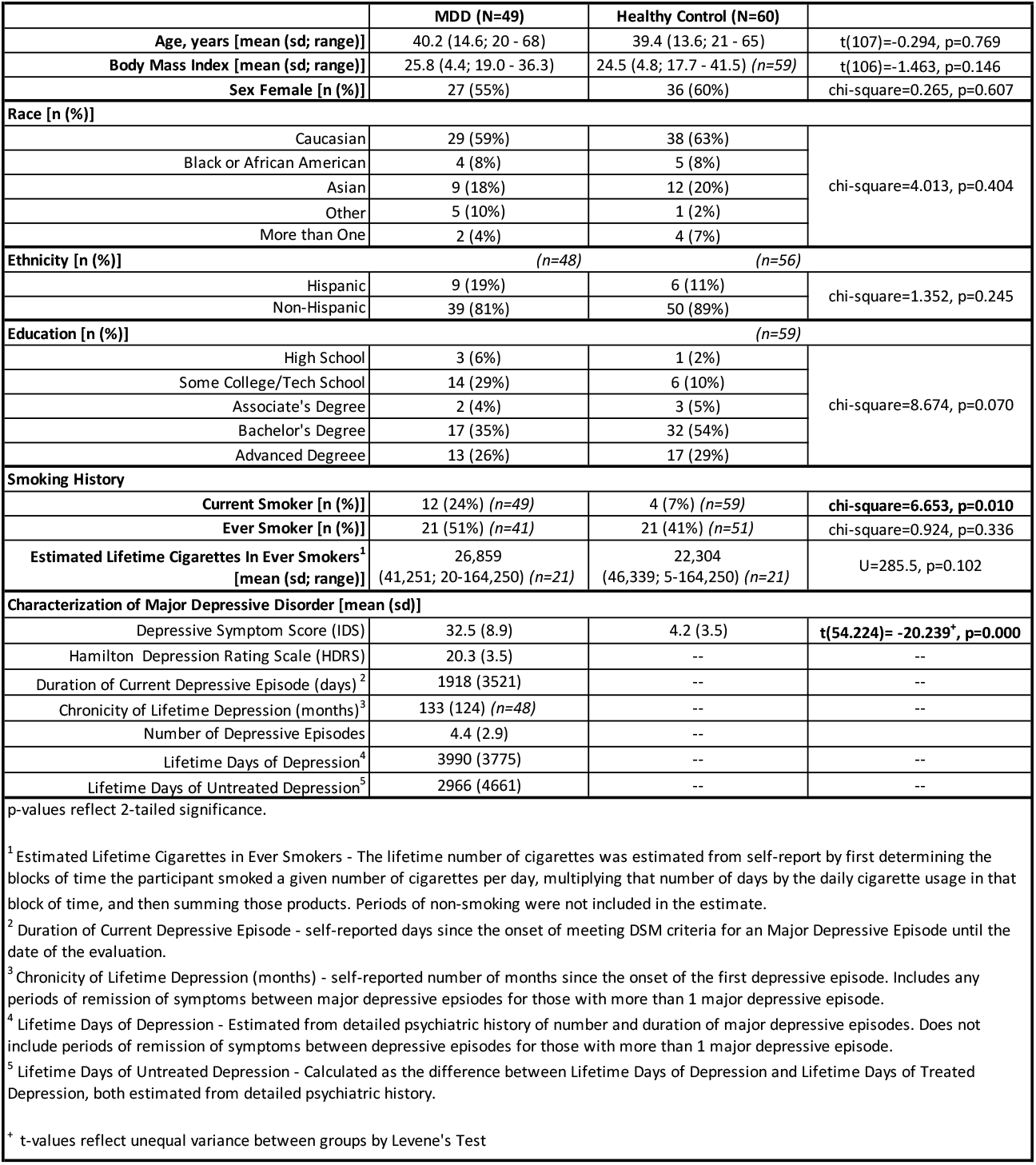
Participant Characterization.

**Table 2:** GrimAge Clock and its Components in MDD and HC.

|  |  |  | Model 1:<sup>1</sup> |  |  | Model 2:<sup>2</sup> |  | Model 3:<sup>3</sup> |  |
| --- | --- | --- | --- | --- | --- | --- | --- | --- | --- |
| MDD vs Control: | MDD: N = 49<br>Mean ± SD | Control: N = 60<br>Mean ± SD | t (df =107) | p-value | Cohen's d | F<sub>MDD</sub><br>(df, df = 1, 105) | p-value | F<sub>MDD</sub><br>(df, df = 1,103) | p-value |
| **Age-Adjusted GrimAge (aka "AgeAccelGrim")** |  |  |  |  |  |  |  |  |  |
| AgeAccelGrim* | 0.30 ± 0.84 | -0.29 ± 1.01 | -3.265 | **0.001** | **0.63** | 7.345 | **0.008** | 6.095 | **0.015** |
| **Age-Adjusted Epigenetic Components of GrimAge** |  |  |  |  |  |  |  |  |  |
| DNAmPACKYRS* | 0.30 ± 0.87 | -0.30 ± 1.00 | -3.306 | **0.001** | 0.64 | 7.042 | **0.009** | 6.668 | **0.011** |
| DNAmCystatinC* | 0.18± 0.84 | -0.23 ± 1.04 | -2.203 | **0.030** | 0.43 | 4.639 | **0.034** | 3.817 | *0.053* |
| DNAmLeptin* | 0.08 ± 1.01 | -0.04 ± 0.96 | -0.599 | 0.551 | 0.12 | 0.235 | 0.629 | 1.421 | 0.236 |
| DNAmTIMP1* | 0.12 ± 0.92 | -0.10 ± 1.04 | -1.133 | 0.260 | 0.22 | 1.812 | 0.181 | 0.951 | 0.332 |
| DNAmADM | -0.11 ± 18.61 | 0.27 ± 15.59 | 0.114 | 0.909 | 0.02 | 0.048 | 0.828 | 0.008 | 0.927 |
| DNAmB2M | 1005.14 ± 51306.85 | -12031.01 ± 69546.19 | -1.125<sup>+</sup> | 0.263 | 0.21 | 1.877 | 0.174 | 1.677 | 0.198 |
| DNAmGDF15 | 3.21 ± 52.28 | -6.86 ± 55.64 | -0.965 | 0.337 | 0.19 | 0.440 | 0.508 | 0.387 | 0.535 |
| DNAmPAI1 | -35.72 ± 1916.48 | -142.78 ± 2646.10 | -0.245<sup>+</sup> | 0.807 | 0.05 | 0.041 | 0.840 | 0.150 | 0.699 |

p-values reflect 2-tailed significance.
All models used age-adjusted metrics of epigenetic age. Age-adjusted metrics were calculated as the residual from regressing GrimAge and its components on chronological age. Age-adjusted GrimAge is denoted as "AgeAccelGrim" to maintain consistency with the literature.
<sup>1</sup> Model 1: Independent samples T-test
<sup>2</sup> Model 2: ANCOVA covaried for current smoking status (binary Y/N)
<sup>3</sup> Model 3: ANCOVA covaried for current smoking status (binary Y/N), sex, and BMI
\* denotes epigenetic age variables that were Blom-transformed to achieve normal distributions.
<sup>+</sup> t-values reflect unequal variance between groups by Levene's Test

**Table 3:** AgeAccelGrim and DNAmPACKYRS Among Current Non-Smokers.

|  |  |  | Model 4:<sup>1</sup> |  |  | Model 5:<sup>2</sup> |  |
| --- | --- | --- | --- | --- | --- | --- | --- |
| MDD vs Control:<br>Non-Smokers | MDD: N = 37<br>Mean ± SD | Control: N = 55<br>Mean ± SD | t (df) | p-value | Cohen's d | F<sub>MDD</sub><br>(df, df = 1, 88) | p-value |
| **Age-Adjusted GrimAge (aka "AgeAccelGrim")** |  |  |  |  |  |  |  |
| AgeAccelGrim* | 0.27 ± 0.77 | -0.18 ± 1.08 | -2.383<sup>+</sup> (89.68) | **0.019** | 0.49 | 3.417 | 0.068 |
| **Age-Adjusted Epigenetic Components of GrimAge** |  |  |  |  |  |  |  |
| DNAmPACKYRS* | 0.31 ± 0.78 | -0.21 ± 1.06 | -2.656 (89.15) | **0.009** | 0.55 | 5.796 | **0.018** |

p-values reflect 2-tailed significance.
All models used age-adjusted metrics of epigenetic age. Age-adjusted metrics were calculated as the residual from regressing GrimAge and its components on chronological age. Age-adjusted GrimAge is denoted as "AgeAccelGrim" to maintain consistency with the literature.
<sup>1</sup> Model 4: Independent samples T-test
<sup>2</sup> Model 5: ANCOVA covaried for sex and BMI
\* denotes epigenetic age variables that were Blom-transformed to achieve normal distributions.
<sup>+</sup> t-values reflect unequal variance between groups by Levene's Test

## Results

### Participant Demographics and Depressive Symptoms

The MDD and control groups did not differ significantly in chronological age, sex distribution, BMI, race, ethnicity, educational attainment, lifetime history of smoking, or estimated number of lifetime cigarettes among smokers (**Table 1**). However, the groups did differ in a binary metric of current tobacco use (Yes/No), with greater current smoking reported in the MDD group (*chi-square*=6.653, *p*=0.010) and, as expected, in scores on the Inventory of Depressive Symptoms (IDS) (*t(54*.*224)*=-*20*.*239, p*=*0*.*000*). Further characterization of participants with MDD is presented in **Table 1**.

### GrimAge Acceleration in Depressed Patients and Healthy Controls

GrimAge was significantly correlated with chronological age in the combined cohort (Spearman *Rho=0*.*968, p<0*.*001)*, and among participants with MDD and controls separately (*MDD: Spearman Rho =0*.*974, p<0*.*001; HC*: Spearman *Rho* =*0*.*961, p*<*0*.*001*) (**Figure 1B**), consistent with strong correlations reported previously(15) and with the inclusion of chronological age as a component of the GrimAge algorithm.

**Figure 1:**
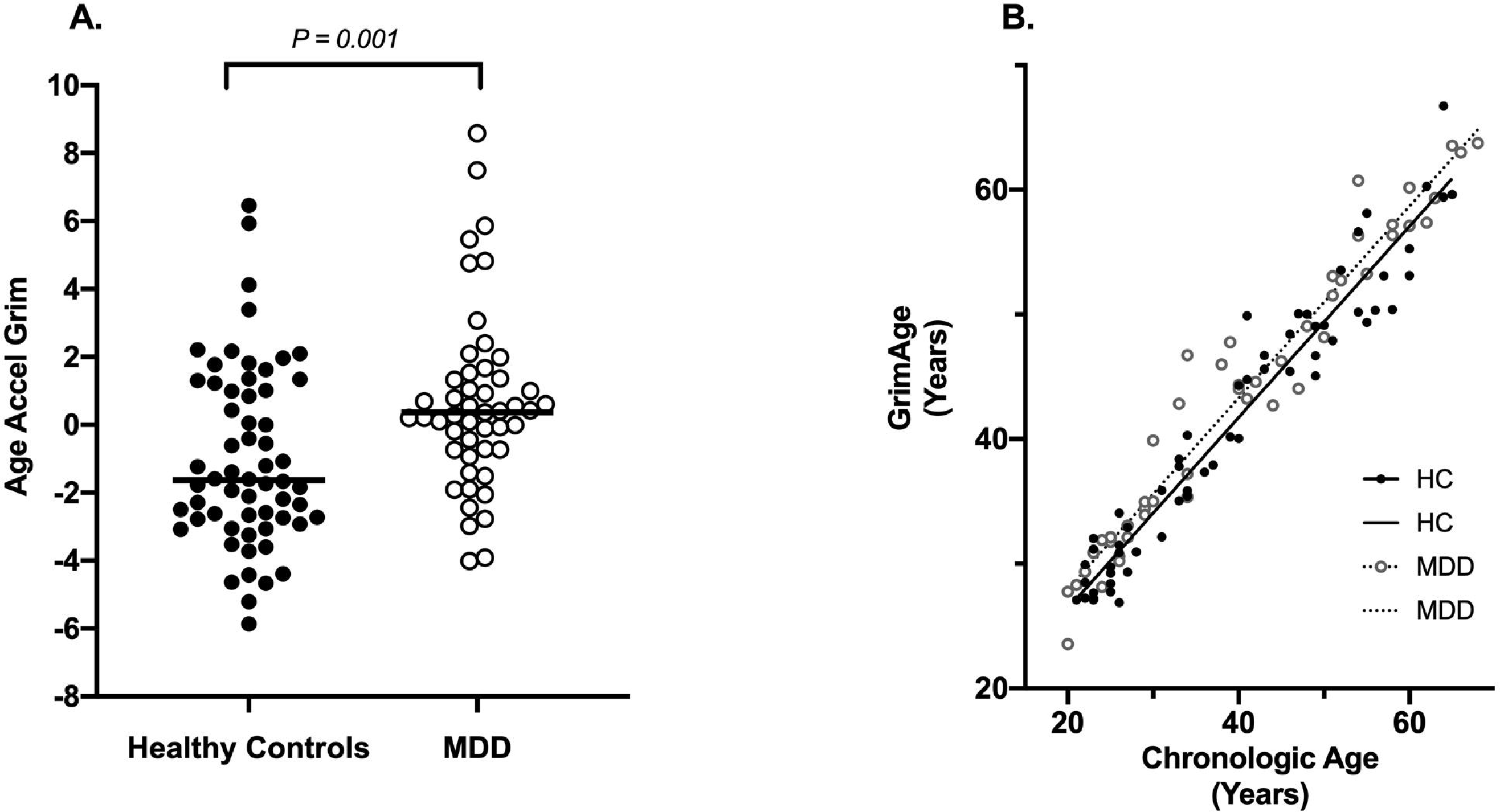
Cross-section differences in AgeAccelGrim between healthy controls and patients with major depressive disorder (MDD). **(A)** Plotted values are raw AgeAccelGrim measures (in years), prior to Blom transformation. P-value reflects two-tailed significance between groups, based on Blom-transformed data to achieve normality of distribution. Horizontal line indicates median AgeAccelGrim within each group. N_healthy control_ = 60, N_MDD_ = 49. (**B**) Plotted values are participants’ chronological age plotted against GrimAge (prior to age-adjustment), demonstrating a strong correlation between chronological age and GrimAge among both participants with MDD and healthy controls (HC) (*Combined: Spearman Rho=0*.*968, p<0*.*001; MDD: Spearman Rho =0*.*974, p<0*.*001; HC: Spearman Rho =0*.*961, p<0*.*001)*.

In our primary analysis, participants with MDD showed significantly greater AgeAccelGrim compared to age-matched healthy controls (*t(107) = -3*.*265, p = 0*.*001; Cohen’s d=0*.*6346*). Based on median values prior to transformation for normality, this equates to 2.0 years of accelerated GrimAge (*MDD: median= 0*.*364 years, interquartile range (IQR)= 2*.*180; HC: median= -1*.*637 years, interquartile range (IQR)=3*.*946)* (**Figure 1A**). The group effect remained statistically significant after adjustment for current smoking (***Model 2:*** *F*_*group*_*(1,109) = 7*.*345, p= 0*.*008)*, and after full adjustment for smoking, sex and BMI (***Model 3:*** *F*_*group*_*(1,103) = 6*.*095, p=0*.*015)* (**Table 2**).

Although AgeAccelGrim was significantly higher in MDDs vs controls, it was not significantly correlated with scores of depression severity on the 17-item Hamilton Depression Rating Scale (HDRS) *(r= -0*.*088, p=0*.*550)* within the MDD group, nor was it associated with reported lifetime days of depression or days of untreated depression *(lifetime days: r= -0*.*009, p= 0*.*953; untreated days: r= 0*.*061, p= 0*.*676)*, chronicity of lifetime depression *(r= -0*.*014, p= 0*.*926)*, or duration of the current depressive episode *(r= -0*.*125, p= 0*.*391)* (See **Table 1** for variable definitions).

### Individual GrimAge Component Markers and the Role of DNAmPACKYRS

Among the benefits of the GrimAge clock is that each of these surrogate DNAm markers can themselves be queried as a means to explore the contribution of each to accelerated cellular aging(15). Of the individual surrogates, only age-adjusted DNAmCystatinC and age-adjusted DNAmPACKYRS significantly differed between MDD and healthy control participants (**Table 2**). The group differences in age-adjusted DNAmCystatinC survived adjustment for smoking, but it missed statistical significance after full adjustment for smoking/sex/BMI. In contrast, the group difference in age-adjusted DNAmPACKYRS survived adjustment for smoking, sex, and BMI (**Table 2**).

Notably, while the DNAmPACKYRS metric was developed as a surrogate for self-reported smoking history(15), the 172 CpGs of which it consists are related to a number of other functions(29,30). To explore whether the DNAmPACKYRS metric captures some underlying biological aspect of aging that is not specific to smoking, we (1) restricted our between-group analysis of AgeAccelGrim and age-adjusted DNAmPACKYRS to current non-smokers in both the MDD and control groups, and (2) covaried for trilevel ‘never,’ ‘former,’ and ‘current’ smoking status in a sub-cohort of participants with detailed data on smoking exposure. Among current non-smokers, we still found a statistically significant difference between the MDD and healthy control groups in both the overall AgeAccelGrim (***Model 4:*** *t(89*.*68)= -2*.*383, p=0*.*019, Cohen’s d=0*.*49)* and in age-adjusted DNAmPACKYRS (***Model 4:*** *t(89*.*15)= -2*.*656, p=0*.*009, Cohen’s d=0*.*55)* (**Table 3**). However, with this smaller sample size, the difference in AgeAccelGrim among non-smokers missed statistical significance after full adjustment for sex and BMI (***Model 5:*** *F*_*group*_*(1,88)=3*.*417, p=0*.*068)*, while the difference in age-adjusted DNAmPACKYRS persisted (***Model 5:*** *F*_*group*_*(1,88)=5*.*796, p=0*.*018)*. Sub-cohort analyses on the effect of lifetime smoking status, as opposed to current smoking status, are further detailed in the **Supplementary Materials**. In brief, differences in both AgeAccelGrim and age-adjusted DNAmPACKYRS between MDD and HC persisted when covarying for trilevel lifetime smoking status (**Supplementary Table 1**).

Of the 172 CpGs included in the DNAmPACKYRS metric, the cg05575921 site, corresponding to the Aryl Hydrocarbon Receptor Repressor (AHRR) gene, was of particular interest in light of recent evidence suggesting a role in post-traumatic stress disorder(31,32). To explore if the cg05575921 methylation site might be contributing to differences in DNAmPACKYRS between MDD and HC independent of smoking exposure, we assessed differences in cg05575921 methylation 1) associated with smoking, and 2) between MDD and HC. These results are presented in the **Supplementary Materials**, but in brief we did not find differences in cg05575921 methylation between MDD and HC. However, we found significant hypomethylation associated with ‘current’ smoking status, consistent with prior reports on the cg05575921 site as a marker for smoking exposure(33–35)(33–35).

## Discussion

In this paper, we present evidence of epigenetic aging in MDD using the recently developed GrimAge metric, which is highly correlated with morbidity and mortality(15), in a unique cohort of somatically healthy participants with severe untreated depression. We found that somatically healthy, unmedicated individuals with MDD exhibit greater GrimAge Acceleration than their healthy normal control counterparts, with a median difference of 2 years of accelerated epigenetic aging. Importantly, the differences in AgeAccelGrim between healthy participants and those with MDD persist after adjustment for smoking, sex, and BMI. This suggests that while these variables contribute to the GrimAge, the association with MDD remains above and beyond what is attributable to them.

To date, there have been very few reports on epigenetic aging in MDD, and none specifically on GrimAge in MDD. The most notable of the reports was from the Netherlands Study of Depression and Anxiety(17). Using an epigenetic age estimate of their own derivation, based on 80 000 CpGs, the group found significant epigenetic age acceleration among depressed patients compared to healthy controls. The effect sizes in both basic and fully adjusted models were small, with Cohen’s d of 0.20 and 0.18, respectively, and showed an average 7.68 months of excess epigenetic aging. In our sample, we found significant differences in GrimAge acceleration between MDD participants and healthy controls, with an effect size of Cohen’s d = 0.63 prior to adjustment and a median excess epigenetic aging of two years. The greater effect size seen here, if replicated, could be due to the specific epigenetic aging metric used or to differences in our sample characteristics.

In light of the significant between-group differences in current tobacco use and in the DNAmPACKYRS component of the AgeAccelGrim metric, it is important to consider the role of tobacco use in explaining our findings, independent of MDD. To do so, we employed two analytic strategies. We (1) restricted our cohort to ‘current non-smokers’, and (2) restricted our cohort to subjects with sufficiently detailed smoking history and adjusted for this history. These analyses were consistent with our hypothesis that both AgeAccelGrim and age-adjusted DNAmPACKYRS are associated with MDD independent of smoking. Our pattern of findings is also consistent with those of Lu et al. in their original publication on GrimAge, where they demonstrated that DNAmPACKYRS predicted lifespan in never smokers(15), suggesting that this composite measure of methylation at 172 CpG sites reflects changes in underlying biological processes not specific to tobacco use. Of these sites, the cg05575921 site and its associated gene – the Aryl Hydrocarbon Receptor Repressor (AHRR) – stand out as potentially relevant to the pathophysiology of psychiatric disease(31,32) and aging(36,37). We compared methylation at this site between MDD and HC and found no significant differences. Nonetheless, in light of evidence for a role the AHRR in atherosclerosis(36,37) and PTSD(31,32) in human studies, in conjunction with evidence from animal studies implicating dysfunction of the AHRR’s target – the Aryl Hydrocarbon Receptor – in aging (notably, brain aging)(38) and neuroinflammation(39), the AHRR remains an interesting and worthwhile target for further research in depression and other psychiatric diseases.

The strengths of our study include the clinically significant degree of current MDD symptomatology of our sample, the verification of MDD diagnosis by structured and independent unstructured psychiatric interviews, and the requirement that all participants remained medication-free for a minimum of 6-weeks prior to entering the study. Despite the severity of their depression, our sample was otherwise somatically healthy, as we excluded individuals with significant current or past history of medical conditions that would likely themselves affect epigenetic aging. As such, we aimed to evaluate the effects of MDD per se, rather than those of comorbid medical conditions or antidepressant use. However, by so doing we may have selected for a sample of patients that is not fully representative of “general community” depression, who may have biological resilience and have thus experienced fewer somatic sequelae of depression. The same is true of our healthy controls, who were also selected for the absence of somatic illnesses and medication. It appears from the raw data (Figure 1) that the AgeAccelGrim difference between healthy controls and MDDs is mainly accounted for by lower-than-expected GrimAge among the HCs, rather than by higher-than-expected GrimAge among MDDs. We interpret this to be due to our careful exclusion of somatic illnesses in both samples, in contrast to the naturalistic community-based samples from which the GrimAge metric was derived.

Limitations of this study include our modest sample size. Our findings therefore require validation in a larger and more diverse cohort. In addition, although participants were largely medication-free for at least 6 weeks prior to enrollment, we cannot rule out lingering effects of prior antidepressant or other medication use. We also had detailed data on lifetime smoking history in only a subset of participants, and therefore could only adjust for current smoking status in our primary analysis. While our sub-cohort analyses demonstrated the effect of MDD on AgeAccelGrim to be robust against adjustment for detailed lifetime smoking exposure, our samples of ‘never’ smokers were too small to achieve adequate power for analysis and our results will require replication in cohorts with extensive data on smoking exposure or within a large cohort of ‘never’ smokers. Finally, while the GrimAge metric was shown to predict mortality in other populations(15), we cannot assert that it indeed does so among individuals with MDD without long-term longitudinal data, or else retrospective data with banked DNA samples.

Individuals with MDD have, on average, lesser life expectancy and greater serious somatic illness comorbidity than non-psychiatrically ill individuals. The reasons for this remain unclear, but our data raise the possibility that the epigenetic changes associated with GrimAge are relevant. However, many questions remain unanswered. Our data do not inform on the stability vs. reversibility of these changes(40,41), on the actual association of these epigenetic changes with later illness and mortality in MDD, on whether any causal relationships exist between MDD and GrimAge, or on whether the GrimAge differences we noted are specific to MDD among other psychiatric illnesses. Furthermore, while our goal was to assess the possibility of accelerated GrimAging in MDD rather than identify its causes, the ultimate aim of this line of research is to identify the biological mechanisms linking MDD to diminished lifespan and “healthspan”(14,15). Advances in understanding the cellular biology underlying serious mental illnesses such as MDD may hold a key to understanding the associated increased risk of illness and death.

## Data Availability

The data that support the findings of this study are available from the corresponding author, EP, upon reasonable request.

## Acknowledgements

The authors gratefully acknowledge the assistance of Kevin Delucchi, PhD, Phuong Hoang, Stacy Ann Miller, the UCSF CTSI Clinical Research Center staff, the PNE Lab research assistants and volunteers, and the research participants.

## Clinical Trial Registration

This clinical study is registered was registered at https://clinicaltrials.gov as trial number NCT00285935.

