## Supplementary Text for "“GrimAge,” an epigenetic predictor of mortality, is accelerated in Major Depressive Disorder"

**SUPPLEMENTARY MATERIAL**

**Covariance for Blood Cell Composition in AgeAccelGrim**

As an additional quality control to ensure that our results were not the result of idiosyncrasies in blood-cell composition, as the GrimAge metric is sensitive to composition, we repeated our analysis of differences in AgeAccelGrim between MDD and HC by covarying for the percentages of monocytes, neutrophils, lymphocytes, eosinophils, and basophils. The significant effect of MDD status persisted when covarying for blood cell composition only (*F_MDD_(1,95) = 12.773, p=0.001*), and with the addition of current smoking status (*F_MDD_(1,94) = 9.117, p=0.003*) and full adjustment for smoking status, sex, and BMI (*F_MDD_(1,92) = 7.794, p=0.006).*

**Sub-Cohort Analyses of Effect of Smoking on Group Differences in AgeAccelGrim**

We repeated our primary analyses of differences in AgeAccelGrim between MDD and HC in a sub-cohort of participants with detailed data available on lifetime smoking history (41 MDD and 51 HC), such that we could covary for tri-level ‘never,’ ‘former,’ or ‘current’ smoking status instead of binary ‘current’ smoking status. The effect in Models 2 and 3 remained significant when analyzed within this subsample (***Model 6*** *(analogous to Model 2)****:*** *F_MDD_(1,89)=8.310, p=0.005;* ***Model 7*** *(analogous to Model 3)****:*** *F_MDD_(1,87)=6.716, p=0.011)* (**Supplementary Table 1)***.*

While it would be desirable to further assess for differences in AgeAccelGrim among ‘never’ smokers, our sample for such an analysis was too small, with only 20 MDD and 30 HC participants classified as ‘never’ smokers, yielding an observed power of only 0.22 after adjustment for sex and BMI.

**Sub-Cohort Analyses of Effect of Smoking on Group Differences in Age-Adjusted DNAmPACKYRS**

We similarly repeated our primary analyses of differences in age-adjusted DNAmPACKYRS between MDD and HC in the same sub-cohort. The group difference in age-adjusted DNAmPACKYRS remained significant when analysis was limited to those with available data on lifetime smoking history and covaried for tri-level smoking history (***Model 6:*** *F_MDD_(1,89)=7.386, p=0.008;* ***Model 7:*** *F_MDD_(1,87)=6.818, p=0.011).* We again did not limit our cohort to ‘never’ smokers, as the sample of 20 MDD and 30 HC yielded inadequate power for this *post-hoc* analysis, with an observed power of only 0.27.

**Effect of Smoking on the cg05575921 Methylation Site, a Component of the DNAmPACKYRS Metric**

Because we identified a statistically significant effect of MDD on the age-adjusted DNAmPACKYRS metric, we considered potential causal mechanisms of the association. One of the methylation sites included in the DNAmPACKYRS metric is the cg05575921 CpG site, which localizes to the Aryl Hydrocarbon Receptor Repressor Gene (AHRR). It has repeatedly been shown that this CpG site demonstrates hypomethylation associated with smoking exposure(1–3), consistent with its selection by machine learning algorithms to the DNAmPACKYRS metric. However, there is also evidence that the AHRR plays a direct causal role in at least some somatic diseases, namely atherosclerosis(4,5), and accumulating evidence for its role in other psychiatric diseases, namely PTSD(6,7). As a result, we sought to assess the relationships between MDD and the cg05575921 methylation site in our cohort.

However, before assessing for disease-specific associations, we first wanted to assess the association of the methylation site with smoking exposure within our sub-cohort with available data on lifetime smoking history (41 MDD and 51 HC). A one-way ANOVA demonstrated a statistically significant effect of lifetime smoking exposure (‘never,’ ‘former,’ ‘current’) on methylation at the cg05575921 site (Brown-Forsythe *F(2,44.68)= 6.374, p=0.004*). Planned contrasts revealed that the effect was largely driven by significant hypomethylation among current smokers (***Never Smokers vs. Any Smoking History:*** *t(49.25)= -2.640, p=0.011;* ***Never vs. Former Smokers:*** *t(42.02)= -0.119, p=0.906;* ***Never vs. Current Smokers:*** *t(17.20)= -3.535, p=0.003;* ***Former vs. Current Smokers:*** *t(27.58)= -2.956, p=0.006).* Among ‘ever’ smokers (defined as the combined ‘former’ and ‘current’ smokers), we also noted a statistically significant negative correlation between the estimated number of lifetime cigarettes and cg05575921 methylation state *(N=42, Spearman Rho= -0.427, p=0.005*). This correlation was at least partially driven by current smokers, as it was attenuated to non-significance when the correlation was restricted to former smokers (*N=28, Spearman Rho= -0.301, p=0.119*). This pattern of findings suggests that, while smoking exposure is associated with hypomethylation at the cg05575921, as has been repeatedly suggested in the literature(1–3), statistically significant differences in methylation state in our study were limited to current smokers. This is in line with a recent paper by McCartney et al., where they examined the epigenetic signatures of smoking in current smokers, and the effects of smoking cessation on DNA methylation in former smokers(8). McCartney et al. report that smoking-related epigenetic changes seemingly require prolonged exposure to cigarette smoke, and are at least partially reversible following cessation(8). To the extent that this is true, our pattern of findings supports the utility of assessing AgeAccelGrim and DNAmPACKYRS among ‘current’ non-smokers when the sample size of ‘never’ smokers is inadequate.

In order to assess the relationship between the AHRR and MDD, as captured by the cg05575921 methylation site, we used an independent samples T-test to assess for differences between MDD and HC in our full cohort and an ANCOVA to assess for differences between MDD and HC after covarying for current smoking status. We did not find statistically significant differences between MDD and HC in cg05575921 methylation state in either the unadjusted or adjusted models.

**Effect of Removing the DNAmPACKYRS and DNAmCystatinC from the GrimAge Algorithm**

To test whether age-adjusted DNAmPACKYRS was driving the significant AgeAccelGrim difference between MDD and controls, we repeated our analysis of group differences in AgeAccelGrim after removing the DNAmPACKYRS component from the GrimAge algorithm as follows: “*AgeAccelGrim_NoDNAmPACKYRS = AgeAccelGrim - 8.3268*0.030398* age-adjusted DNAmPACKYRS.”* This method has been previously described for the same purpose(9). Using this new metric (“AgeAccelGrim_NoDNAmPACKYRS”), the difference between MDD and healthy control participants was attenuated to non-significance *(****Model 1:*** *t(107)=* *-1.534, p=0.128, Cohen’s d=0.30).* Adjusting for smoking, sex and BMI further attenuated any group differences *(****Model 2****: F_MDD_(1,105)=* *2.161, p=0.145;* ***Model 3:*** *F_MDD_(1,103)=* *1.071, p=0.303).* The same method was applied to subtract DNAmCystatinC from the GrimAge algorithm (*“AgeAccelGrim_NoCystatinC = AgeAccelGrim – 8.3268*3.5E-6*age-adjusted DNAmCystatinC)*, and the group effect persisted in all models. However, it should be noted that the DNAmPACKYRS component of GrimAge is weighted much more heavily within the algorithm than DNAmCystatinC, precluding direct comparison.
