## Supplementary Table 1 for "“GrimAge,” an epigenetic predictor of mortality, is accelerated in Major Depressive Disorder"

**Supplementary Table 1: Sub-Cohort Analyses of Effect of Smoking on GrimAge and DNAmPACKYRS**

| | Model 6: [1] | | Model 7: [2] | |
|---|---|---|---|---|
| **MDD vs Control:** | **$F_{MDD}$ (df, df = 1, 89)** | **p-value** | **$F_{MDD}$ (df, df = 1, 87)** | **p-value** |
| Age-Adjusted GrimAge* (aka "AgeAccelGrim") | 8.310 | **0.005** | 6.716 | **0.011** |
| Age-Adjusted DNAmPACKYRS* | 7.386 | **0.008** | 6.818 | **0.011** |

p-values reflect 2-tailed significance.
All models used age-adjusted metrics of epigenetic age. Age-adjusted metrics were calculated as the residual from regressing GrimAge and its components on chronological age. Age-adjusted GrimAge is denoted as "AgeAccelGrim" to maintain consistency with the literature.

[1] Model 6: ANCOVA covaried for trilevel smoking status (Never, Former, Current) ($N_{MDD}$ = 41, $N_{HC}$ = 51)

[2] Model 7: ANCOVA covaried for trilevel smoking status (Never, Former, Current), sex, and BMI ($N_{MDD}$ = 41, $N_{HC}$ = 51)

* denotes epigenetic age variables that were Blom-transformed to achieve normal distributions.
